# Clinical epidemiology of easily recognisable congenital malformations in Gambian newborns: A descriptive cohort study embedded in a clinical trial

**DOI:** 10.1101/2025.11.03.25339407

**Authors:** Shashu Graves, Nathalie Beloum, Bully Camara, Usman N. Nakakana, Madikoi Danso, Fatoumata Sillah, Joquina C. Jones, Edrissa Sabally, Siaka Badjie, Omar Jarra, Sulayman Bah, Abdoulie Suso, Ebrahim Ndure, Yusupha Njie, Christian Bottomley, Halidou Tinto, Umberto D’Alessandro, Helen Brotherton, Anna Roca

## Abstract

**Background:** The prevalence of congenital malformations remains high and significantly contributes to neonatal morbidity and mortality in West Africa. However, there is a gap in the existing literature regarding the clinical epidemiology of congenital malformations in the African subregion. This study aimed to describe the clinical presentation, prevalence, risk factors, and adverse neonatal outcomes of newborns with easily recognisable congenital malformations in urban Gambia.

**Methods:** This descriptive cohort study consisted of secondary analyses of a clinical trial data (PregnAnZi2 trial) for liveborn neonates delivered between October 2017 to May 2021 at two public health facilities in The Gambia. Congenital malformations were detected by clinical examination at birth with active and passive surveillance for 28 days after delivery. Congenital malformations were classified according to ICD11 definitions, with sub-classification into major or minor malformations as per WHO guidance.

**Results:** 140 out of 6750 neonates (2.1%) had at least one congenital malformation, with 29.3% (44/150) of these malformations classified as major. The musculoskeletal system was most frequently affected (58%, 87/150) with talipes equinovarus as the most common major malformation identified (43%, 19/44). A history of previous miscarriage was associated with an increased risk of congenital malformations (aOR 1.72, 95% CI 1.02 to 2.73, p value=0.04). Newborns with a congenital malformation were more likely to experience adverse neonatal outcomes than newborns without, with higher odds of low 1 minute Apgar score, (cOR 3.09, 95% CI 1.62 to 5.48, p <0.001), low 5 min Apgar score (cOR 6.10, 95% CI 2.10 to 14.62, p <0.001), hospitalisation during the neonatal period (cOR 4.42, 95% CI 2.77 to 6.83, p <0.001) and mortality within 24hrs of delivery (cOR 48.75, 95% CI 11.08 to 215.62, p <0.001). Among 70 neonatal deaths recorded by day 28 after birth, 15 (21.4%) were newborns with congenital malformation. The circulatory and skeletal systems were equally affected among congenital malformations recorded in neonatal deaths by day 28 (23.8%, 5/21).

**Conclusion:** Despite a relatively low prevalence, easily recognisable congenital malformations are associated with substantial neonatal morbidity and mortality. Improved antenatal diagnosis of congenital malformations is urgently required to optimise delivery strategies and improve perinatal outcomes, especially for women experiencing previous miscarriage.

**Trial Registration:** NCT03199547: Clinicaltrials.gov. Registered on 23rd June 2017

## Introduction

Globally, congenital malformations affect approximately 6% of newborns, representing more than 8 million babies annually (1, 2). Among these cases, major malformations, which involve a structural change with significant medical, social, or cosmetic consequences or require medical or surgical intervention (3), occur in 2-3 % of live births and contribute to 20-30% of stillbirths (4). However, there is a significant geographical variation in the prevalence of congenital malformations, with 94% of newborns with a major congenital malformation in low- and middle-income countries (1, 5).

As global neonatal mortality rates decline, the proportion of neonatal deaths attributed to congenital malformations has increased (6). Congenital malformations account for 6% of neonatal deaths (6), with 95% of these deaths occurring in low- and middle-income countries (1, 5). For those surviving the neonatal period, congenital malformations can lead to life-long consequences, including chronic illness, long-term disability, and premature death. They account for 25 million disability-adjusted life years (DALYs) globally and have a substantial impact on lost life years (LLYs) (1, 7–10).

Congenital malformations can be classified according to the underlying developmental mechanism. About one-third of congenital malformations might have a genetic cause, with 17% related to single-gene abnormalities and 10% to chromosomal changes (11). Another ∼10% are associated with environmental and maternal factors such as exposure to congenital infections or harmful substances, maternal infections (e.g. rubella) or chronic maternal diseases (e.g. diabetes) (12) which can impact intra-uterine growth and post-conception development (10) or maternal nutritional deficiencies. The remaining ∼60% are of unknown aetiology. Part of the reason so many are of unknown aetiology is that congenital malformations often result from multifactorial causes involving complex interactions between undefined genetic variants and the environment (1, 12). Given this complexity, the most comprehensive and widely used classification is the system-based approach to diagnosis as per the recently updated International Classification of Diseases 11^th^ Revision (ICD11) (13).

Globally recognized risk factors for congenital malformations include a family history, parental consanguinity, and advanced maternal age (6, 14). However, context-specific risk factors for congenital malformations among African newborns remain inadequately described, contributing to a gap in the evidence. Key risk factors may include antenatal and perinatal maternal health (such as limited access to antenatal care), maternal nutritional deficiencies (such as in iodine or folate) (15), maternal exposure to infectious agents including syphilis, rubella, Cytomegalovirus, Zika virus, exposure to teratogens (such as pesticides, herbicides, traditional medicines with unknown chemical constituents). Unregulated medicine use, including antibiotics, also poses a potential risk. Other considerations involve exposure to surface water pollution as a source of drinking water or exposure to hazardous waste (16). The relative contributions of these risk factors may vary by sub-region and season (17).

Robust data on the burden of congenital malformations are lacking in the West Africa sub-region where the neonatal mortality rate is the highest. These data are required to inform the implementation of preventive public health policies aimed at reducing the prevalence of congenital malformations by targeting specific risk factors in the region. In The Gambia, a small country in west Africa, the reported prevalence of congenital malformations was 62.5 per 1000 births in 2006 (1). A study based on a relatively small cohort indicated that major congenital malformations contributed to 13% of neonatal hospitalisations and up to 8% of neonatal deaths in an urban health facility (18). This study aims to investigate the clinical epidemiology of easily recognisable congenital malformations in a large cohort of Gambian newborns delivered in health facilities and following a relatively healthy pregnancy. The objectives include determining the prevalence, describing the spectrum of easily recognisable congenital malformations, identifying risk factors associated with the development of malformations, and estimating the impact of malformations on adverse perinatal outcomes, with stratification by severity (major versus minor).

## Methods

### Study Design

This analysis is based on data collected as part of the PregnAnZI-2 clinical trial, a phase-III, double-blind, placebo-controlled trial (clinicaltrials.gov NCT03199547) conducted from October 2017 to May 2021 (19). In this trial, nearly 12,000 pregnant women in The Gambia and Burkina Faso received intrapartum oral azithromycin or placebo, to assess effectiveness on a composite primary outcome of neonatal sepsis or mortality (20). As the trial intervention was given during labour, it could not have impacted on the aetiology or rates of congenital malformations, hence participants from both trial arms were included in this study. The analysis reported here is restricted to the Gambian cohort of women and their offspring.

### Study Setting

The Gambia is the smallest country in West Africa with population of about 2.6 million people in 2020, of whom approximately 570,000 were women of reproductive age, and with a birth rate of 33.7/1000 inhabitants (21). In 2020, the neonatal mortality rate was 25.7 deaths per 1000 live births (22). Participants were enrolled at two government-run health facilities located in the Gambia’s urban region; Serekunda Health Centre (SHC), which manages approximately 2,000 deliveries per year; and Bundung Maternal and Child Health Hospital (BMCHH) which serves a larger catchment area of ∼50,000 inhabitants and manages ∼5,000 deliveries per year (19). In both health facilities, antenatal clinics are scheduled monthly; each antenatal clinic includes a health talk, a thorough assessment of pregnant women involving history-taking, physical examinations, and relevant laboratory tests as needed. Additionally, the session includes the administration of tetanus toxoid immunizations and the provision of iron/folate supplementation (23). At the time of the PregnAnZI-2 trial data collection, SHC lacked the capacity to provide emergency obstetric care, including surgical care; and lacked a neonatal unit. Thus, women in need of an emergency caesarean section were referred to Kanifing General Hospital, 3.4km away from SHC. Neonates born at SHC and requiring hospital care were transferred to the neonatal unit at Kanifing General Hospital or to BMCHH. BMCHH provided emergency obstetric and neonatal care and on rare occasions transferred women or neonates to the national referral centre, Edward Francis Small Teaching Hospital in Banjul (EFSTH), (15.5Km away from BMCHH) which has paediatric surgical services and a level 2+ neonatal unit.

The three largest ethnic groups in this Gambian region are Mandinka, Fula and Wolof. Marriage between relatives is common, with 30% of marriages being between first cousins (24). The climate is typical of that in the Sub-Sahel region, with a short rainy season from June to October and longer dry season from November to May (19).

### Participants, recruitment, and follow-up

The PregnAnZI-2 trial consented pregnant women aged ≥16 years during antenatal care at the study health facilities, with enrolment during labour. Exclusion criteria included women with known HIV infection, any chronic or acute conditions, planned caesarean section or known required referral, known severe congenital malformation, intrauterine death or allergy to macrolides, and drugs known to prolong QT interval taken during the last 2 weeks, such as chloroquine, quinine, piperaquine, and erythromycin (19). In the trial we did not report any participant excluded following a known severe congenital malformation before delivery, likely due to the lack of routine detailed antenatal scans in the Gambia health facilities. This secondary analysis included all women recruited in The Gambia who delivered a live-born newborn at the trial health facilities.

### Study procedures

Written informed consent to participate in the trial was obtained during antenatal visits and verbally confirmed during labour. Women’s socio-demographic and perinatal clinical data were transcribed from the maternal antenatal card and partograph to an electronic case report form (eCRF) by research nurses before discharge from the health facility.

Between 4 and 24h after birth and before discharge, a detailed clinical examination was carried out by either a trained research clinician or paediatrician, including the newborn’s survival status at the time of assessment (alive or dead), and documentation of any easily recognisable congenital malformation. Newborns who were hospitalised immediately after birth had the same examination in the neonatal ward by the research clinicians. However, for those who died immediately after birth (not seen by research clinicians), the documentation on easily recognisable congenital malformations was based only on the report from the trial nurses present at the time of delivery; this data is not part of this study. Congenital malformations included in this study were those diagnosed by a team of six research clinicians, including five trained medical doctors and one paediatrician. The research clinicians were responsible for assessing all the trial newborns before discharge and diagnosing easily recognizable congenital malformations. Each medical doctor had received formal training in clinical examination and was experienced in assessing newborns. The paediatrician, who had specialized training and experience in paediatric care, provided additional expertise and support. In cases where a medical doctor had doubts regarding the diagnosis of a congenital malformation, they consulted the paediatrician for confirmation. This collaborative approach helped ensure accurate diagnoses and reliable data collection. Both the medical doctors and the paediatrician were tasked with documenting the presence or absence of congenital malformations as part of the study. Detailed investigations such as echocardiography, neuroimaging, abdominal ultrasound, and genetic testing were not routinely available. Congenital malformations requiring urgent surgical intervention were referred to the National Neonatal Referral Hospital (EFSTH). Newborns with congenital malformations needing immediate medical attention were admitted to either BMCHH or EFSTH, where they received further management from paediatricians and neonatologists. These referral centres provided ongoing clinical monitoring and appropriate care for affected newborns. Admissions and referrals to the BMCHH or EFSTH neonatal unit were based on either the research clinicians’ or health facility doctors’ assessment of clinical status.

From discharge to the end of the neonatal period (28 days), participants were followed by passive case detection. The study team actively encouraged mothers to attend the health facilities anytime they or their infant were unwell. Active follow-up of both mother and newborn were conducted by study nurses in the form of home visits on day 28 (+/- 4 days). Newborns requiring further examination by the trial clinicians were referred to the trial sites. The trial clinician or a paediatrician then assessed the mother/newborn and decided whether to admit them for further investigations and management (25). Written serious adverse events reports were filled for any newborn requiring hospitalisation and management, including those with a congenital malformation. In the event of death, the report included the cause of death based on the trial clinicians’ assessment using clinical notes and results of investigations.

### Outcome and variables of interest

The primary outcome was the occurrence of congenital malformations among live births as detected by the research clinician or paediatrician on clinical examination during the discharge review. Congenital malformations were defined as structural anomalies that were easily recognisable on physical examination. The congenital malformations were categorised and coded by organ-system as per the ICD11 classification (13) by an experienced neonatologist. The severity of congenital malformation was classed as major if it was a structural change with significant medical, social or cosmetic consequences or required medical or surgical intervention, in keeping with WHO guidance (3). Minor malformations were defined as structural changes posing no significant health problem in the neonatal period and with limited social or cosmetic consequence (3). Multiple malformations occurring in the same patient were coded separately during descriptive analyses with further classification as major congenital malformation if one or more major congenital malformations was present. To explore the aetiological factors associated with congenital malformation, we developed a conceptual framework based on existing evidence and biological plausibility (Figure 1). Variables were divided into pre-natal, peri-conception and antenatal exposures, with intersectional relationships described. The conceptual framework also considers potential adverse perinatal outcomes associated with congenital malformations. It guided the choice of variables for the logistic regression analysis (see details below in Statistical analysis).

**Figure 1:**
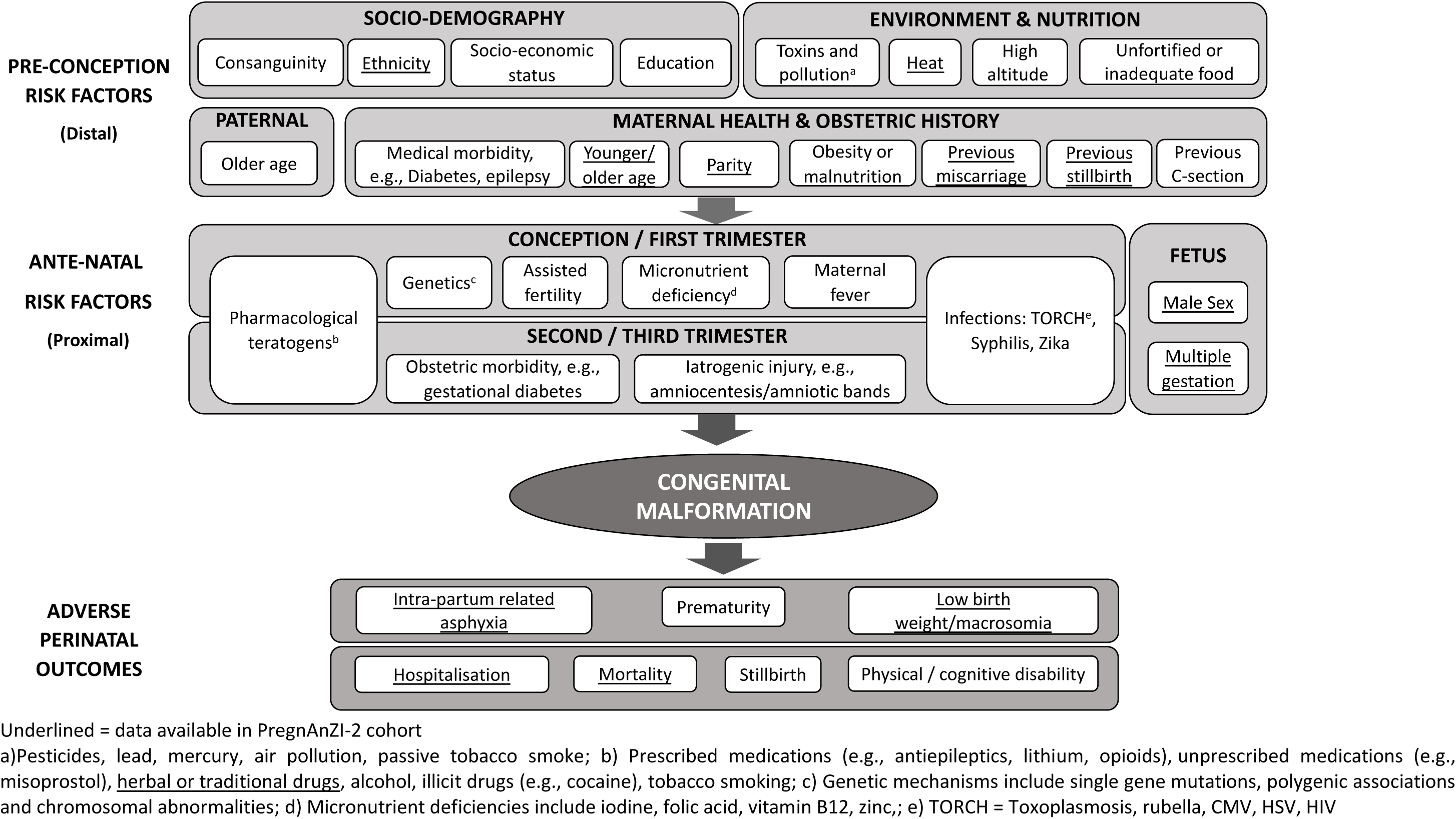
Conceptual Framework to understand risk factors and outcomes of congenital malformations

### Operational definition

The term “Relatively healthy pregnant women” used in our study refers to pregnant women in low-resource health systems such as The Gambia, where antenatal detection of high-risk women/neonates is challenging; this population is representative of pregnant women thought to be at low risk at the time of enrolment. Our study was embedded into a clinical trial that excluded “known” high-risk pregnancies (acute/chronic condition, planned caesarean section, severe congenital malformation, foetal macrosomia, etc.); in practice, often high-risk pregnancies were undiagnosed and were therefore included in the trial due to the lack of rigorous antenatal diagnosis in the trial health facilities.

### Data management and statistical analyses

Research Electronic Data Capture (RedCAP) was used for data capture with consistency checks, data validation and cleaning carried out at regular intervals to ensure high data quality. The data was pseudoanonymised, with each participant assigned a random number. R (version 4.3.1) was used for all analyses. An individual-level analysis was conducted where newborns were classified according to the presence/absence of congenital malformation. Those classified as positive were further classified as positive for a major congenital malformation if one or major congenital malformations was recorded. For analyses describing the distribution of different types of CM, we used the denominator “total number of different occurrences of CM.” In contrast, for analyses involving neonates diagnosed with CM, the denominator “total number of neonates with CM” was used. Crude odds ratios (OR), 95% confidence intervals (CI), and p-values were calculated in unadjusted and adjusted analysis to explore associations between congenital malformation and socio-demographic, environmental, and obstetric factors. An adjusted logistic regression model was developed for each independent variable with a p-value<0.2 for any categorical values in the unadjusted regression. Based on the literature and the data available in our dataset, factors considered to be potential confounders (e.g., maternal age) were included in the adjusted analysis. Statistical significance for the adjusted analysis was pre- determined at p-value <0.05.

## Results

### Baseline characteristics

Overall, 6,750 live births were included in the study (Figure 2), of whom 51.7% (3491/6750) were male, with median birth weight of 3.1Kg (interquartile range (IQR) of 2.8-3.4) and predominantly singletons (97.3%, 6571/6750). Out of all live-born neonates, 140 were diagnosed with at least one easily recognizable congenital malformation, resulting in a prevalence of 2.1% (140/6750); among them, 27.9% (39/140) had at least one major easily recognisable congenital malformation (Figure 2). Multiple malformations were present in 4.3% (6/140) of affected newborns (Table 1).

**Figure 2:**
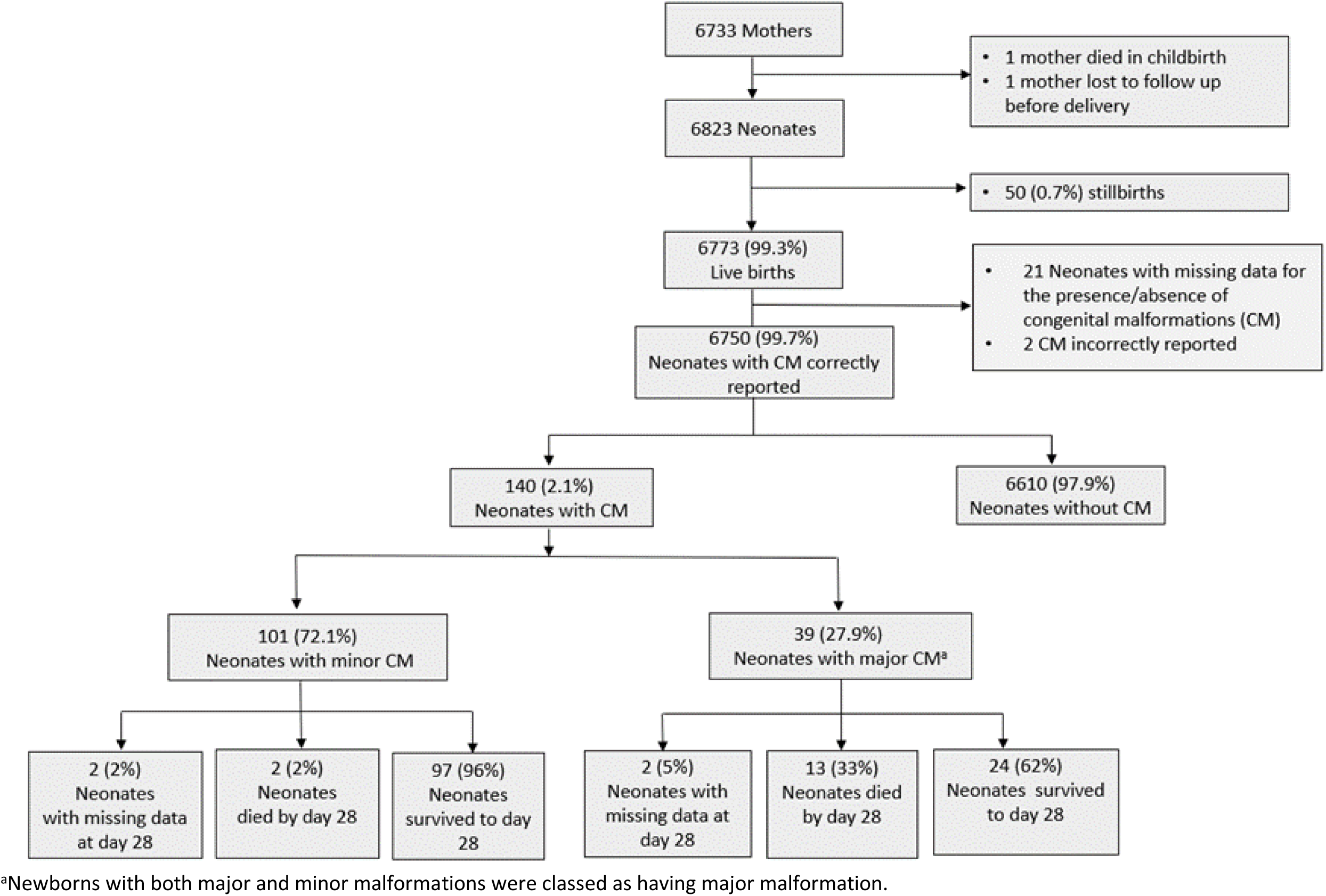
Study Profile for easily recognisable congenital malformations among live-born Gambian neonates following relatively healthy pregnancy

**Table 1:**
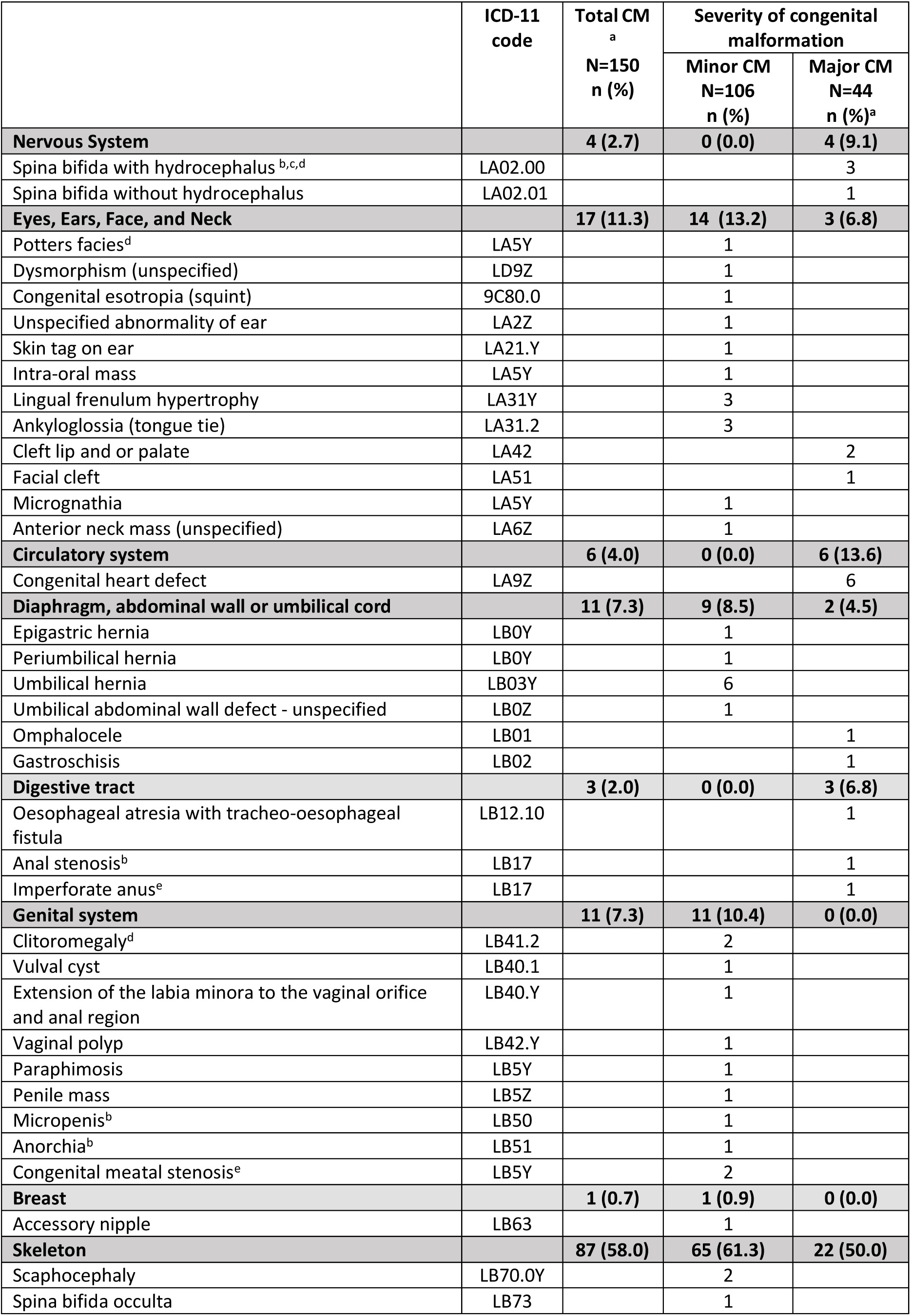
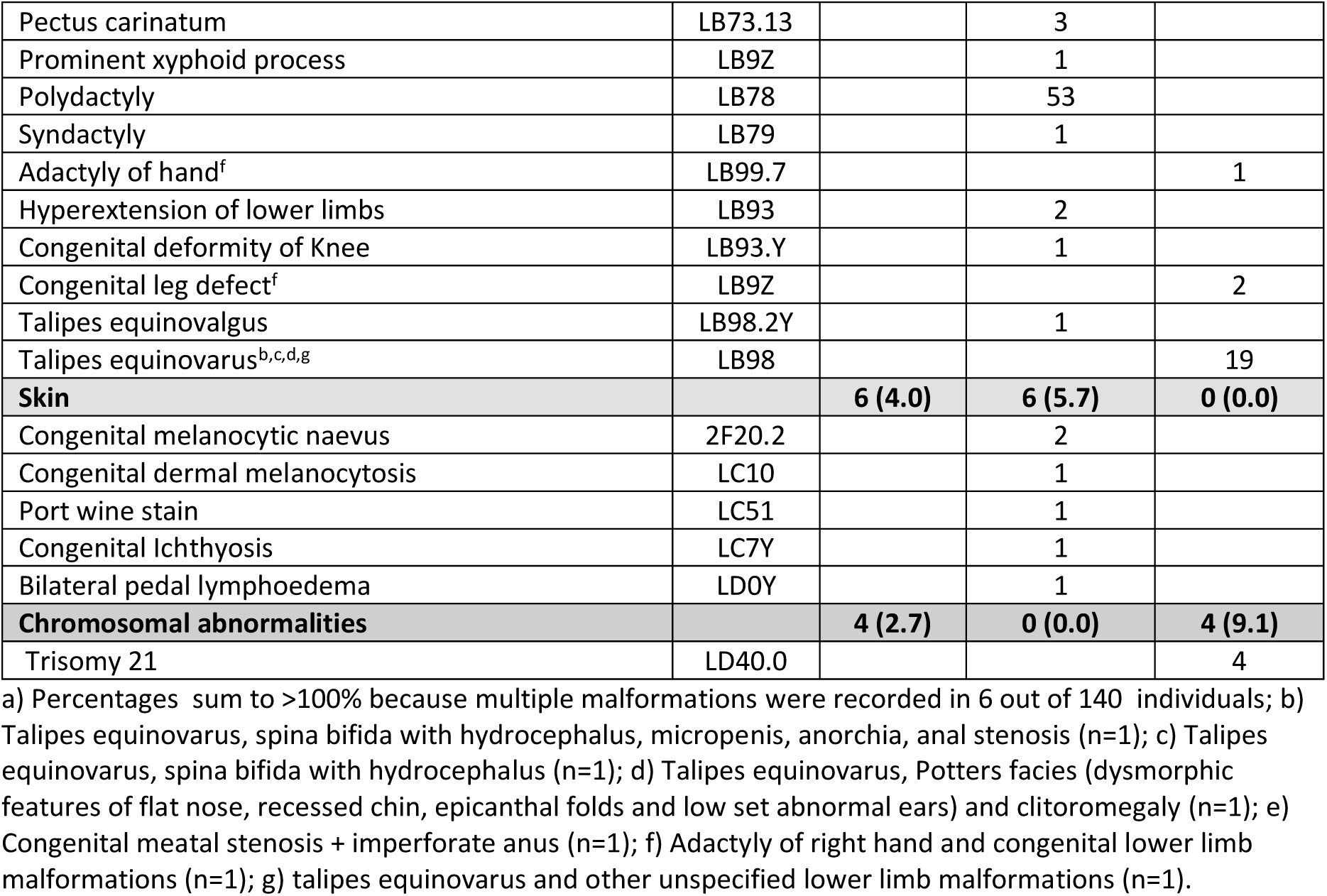
Easily recognisable congenital malformations in a cohort of Gambian neonates born following relatively healthy pregnancy, classified and coded as per ICD-11 (13)

### Types of easily recognisable congenital malformations

In total, 150 different occurrences of easily recognisable congenital malformations were recorded. Nearly one-third (29.3%, 44/150) of them were classified as major (Figure 2, Table 1), with the musculoskeletal system comprising the largest proportion of easily recognisable congenital malformations (58.0%, 87/150) (Table 1), (Figure 3). Talipes equinovarus was the most frequently observed major malformation (43%, 19/44) followed by suspected congenital heart defects (13.6%, 6/44) and spina bifida with or without congenital hydrocephalus (9.1%, 4/44)(Table 1). There were 38 distinct minor malformations, with polydactyly the most common (50.0%, 53/106) (Table 1), (Figure 3).

**Figure 3:**
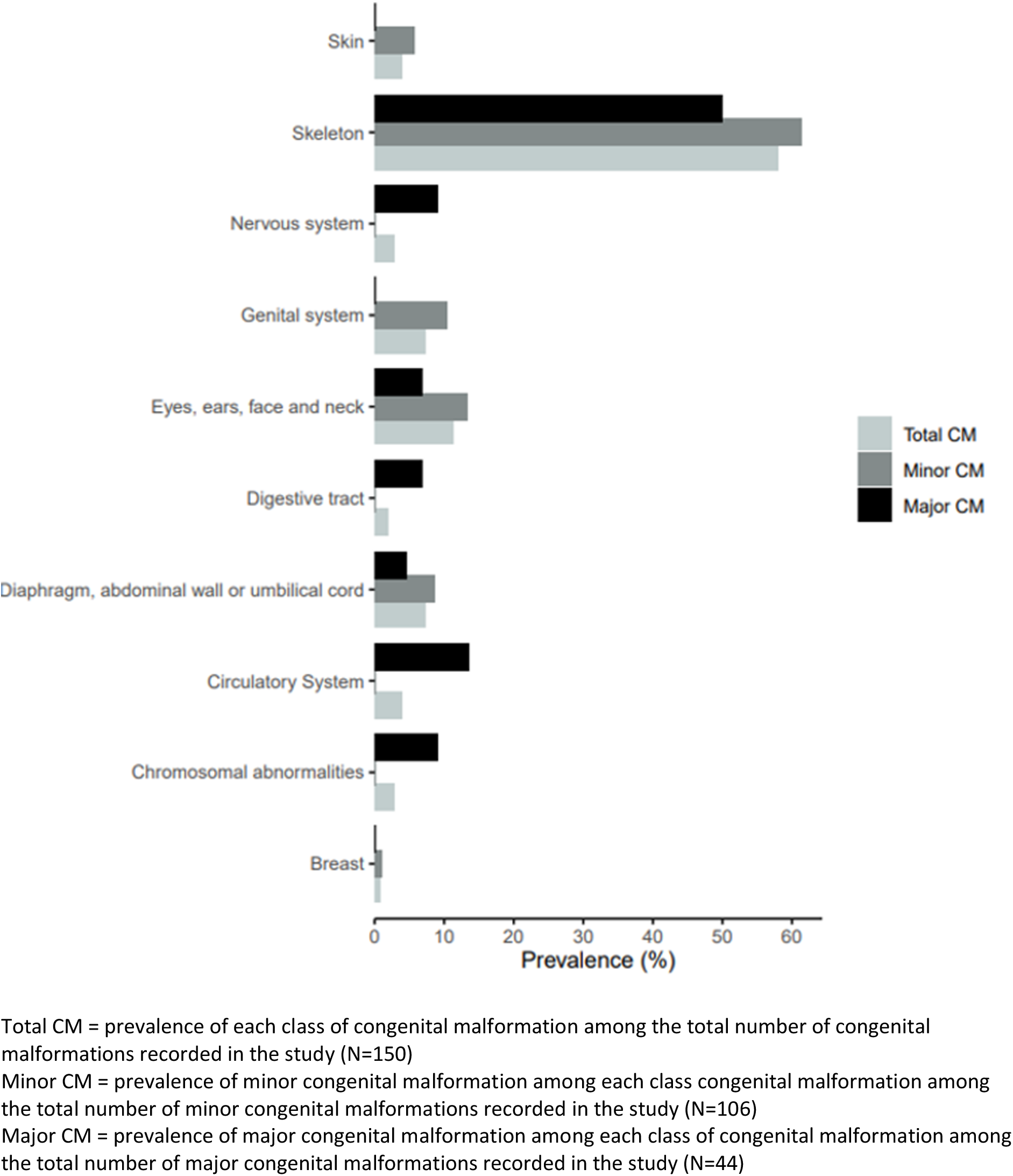
Prevalence of easily recognisable congenital malformations among Gambian neonates by type and severity of the malformation classified as per ICD-11

### Risk factors for easily recognisable congenital malformations

A history of previous miscarriage was associated with increased odds of the newborn having a congenital malformation (OR:1.70, 95%CI:1.00-2.77, p-value=0.03). No other risk factors were significantly associated with malformations (Table 2).

**Table 2:** Maternal, prenatal, and antenatal factors associated with easily recognisable congenital malformations in a cohort of Gambian neonates born following relatively healthy pregnancy

|  | Total<br>n (%)<br>N=6750 | Congenital Malformations |  | Unadjusted |  | Adjusted |  |
| --- | --- | --- | --- | --- | --- | --- | --- |
|  |  | Yes, n (%)<br>n=140 | No, n (%)<br>n=6610 | cOR (95%CI) | p-value | aOR (95%CI) | p-value |
| Maternal factors |  |  |  |  |  |  |  |
| Age (Years) |  |  |  |  | 0.30 |  | 0.31 |
| Median (IQR) | 27 (23-31) |  |  |  |  |  |  |
| 20-35 | 5453 (80.8) | 106 (1.9) | 5347 (98.1) | 1 |  | 1 |  |
| 16-19 | 632 (9.4) | 17 (2.7) | 615 (97.3) | 1.39 (0.80-2.27) |  | 1.24 (0.71-2.04) |  |
| >35 | 665 (9.8) | 17 (2.6) | 648 (97.4) | 1.32 (0.76-2.16) |  | 1.46 (0.84-2.39) |  |
| Mother's Ethnicity |  |  |  |  | 0.45 |  |  |
| Mandinka | 2701 (40.0) | 57 (2.1) | 2644 (97.9) | 1 |  |  |  |
| Fula | 1212 (18.0) | 28 (2.3) | 1184 (97.7) | 1.10 (0.69-1.72) |  |  |  |
| Wolof | 1040 (15.4) | 17 (1.6) | 1023 (98.4) | 0.77 (0.43-1.30) |  |  |  |
| Jola | 870 (12.9) | 23 (2.6) | 847 (97.4) | 1.26 (0.76-2.03) |  |  |  |
| Others | 927 (13.7) | 15 (1.6) | 912 (98.4) | 0.76 (0.41-1.32) |  |  |  |
| Father's Ethnicity<sup>a</sup> |  |  |  |  | 0.36 |  |  |
| Mandinka | 2611 (38.7) | 60 (2.3) | 2551 (97.7) | 1 |  |  |  |
| Fula | 1158 (17.1) | 23 (2.0) | 1135 (98.0) | 0.86 (0.52-1.38) |  |  |  |
| Wolof | 1165 (17.3) | 23 (2.0) | 1142 (98.0) | 0.86 (0.52-1.37) |  |  |  |
| Jola | 830 (12.3) | 21 (2.5) | 809 (97.5) | 1.10 (0.65-1.80) |  |  |  |
| Others | 985 (14.6) | 13 (1.3) | 972 (98.7) | 0.57 (0.30-1.01) |  |  |  |
| Parity |  |  |  |  | 0.36 |  |  |
| Para >=4 | 2513 (37.3) | 55 (2.2) | 2458 (97.8) | 1.05 (0.67-1.68) |  |  |  |
| Primiparous | 1680 (24.9) | 37 (2.2) | 1643 (97.8) | 1.06 (0.65-1.74) |  |  |  |
| Para 2 | 1393 (20.6) | 29 (2.1) | 1364 (97.9) | 1 |  |  |  |
| Para 3 | 1164 (17.2) | 19 (1.6) | 1145 (98.4) | 0.78 (0.43-1.39) |  |  |  |
| Season of delivery |  |  |  |  | 0.34 |  |  |
| Dry | 4568 (67.7) | 100 (2.2) | 4468 (97.8) | 1 |  |  |  |
| Rainy | 2182 (32.3) | 40 (1.8) | 2142 (98.2) | 0.83 (0.56-1.22) |  |  |  |
| Previous stillbirths<sup>b</sup> |  |  |  |  | 0.66 |  |  |
| No | 6636 (98.3) | 137 (2.1) | 6499 (97.9) | 1 |  |  |  |
| Yes | 113 (1.7) | 3 (2.7) | 110 (97.3) | 1.29 (0.26-3.96) |  |  |  |
| Previous miscarriage<sup>c</sup> |  |  |  |  | 0.03 |  | 0.04 |
| No | 6138 (91.0) | 120 (2.0) | 6018 (98.0) | 1 |  | 1 |  |
| Yes | 610 (9.0) | 20 (3.3) | 590 (96.7) | 1.70 (1.00-2.77) |  | 1.72 (1.02-2.37) |  |
| Previous intake of traditional medicine<sup>d</sup> |  |  |  |  | 0.88 |  |  |
| No | 5573 (82.6) | 115 (2.1) | 5458 (97.9) | 1 |  |  |  |
| Yes | 1173 (17.4) | 25 (2.1) | 1148 (97.9) | 1.03 (0.64-1.61) |  |  |  |
| Neonatal factors |  |  |  |  |  |  |  |
| Sex |  |  |  |  | 0.34 |  |  |
| Female | 3259 (48.3) | 62 (1.9) | 3197 (98.1) | 1 |  |  |  |
| Male | 3491 (51.7) | 78 (2.2) | 3413 (97.8) | 1.18 (0.83-1.68) |  |  |  |
| Plurality of birth |  |  |  |  |  |  |  |
| Singleton | 6571 (97.3) | 140 (2.1) | 6431 (97.9) | - | - |  |  |
| Twin | 179 (2.7) | 0 (0.0) | 179 (100.0) |  |  |  |  |

a) Ethnicity father missing in n=1; b) History of stillbirths missing in n=1; c) History of miscarriage missing in n=2; d) Previous intake of traditional medicine missing in n=4

### Adverse perinatal outcomes

Newborns with an easily recognisable congenital malformation were more likely to have an adverse perinatal outcome compared to those without (Figure 3, Table 3). Higher odds of both low 1-minute Apgar score (OR:3.09, 95%CI:1.62-5.48, p-value<0.001) and low 5-minute Apgar score (OR:6.10, 95%CI:2.10-14.62, p-value<0.001) were observed. The odds of being admitted to hospital were more than four-fold higher than in newborns with no malformation (OR:4.42, 95%CI:2.77-6.88, p-value<0.001); all mortality outcomes were higher, especially death within 24hrs of delivery (OR:48.75, 95%CI:11.08-215.62, p-value<0.001)(Table3, Figure 3). Irrespectively of the severity of congenital malformations (minor or major), hospitalisation during the neonatal period was higher among newborns with congenital malformations than in those without (table 4). In total, 15 out of 70 neonatal deaths (21.4%) occurred in newborns with congenital malformations and 50% (5 out of 10) of deaths within the first 24 hours after birth were among newborns with congenital malformations (table 4); these early and overall neonatal deaths were more common among those with major congenital malformations (Figure 4). Among neonates with congenital malformations, those affecting the skeleton were most frequently recorded among admitted neonates. Similarly, skeletal, circulatory system and chromosomal congenital malformations were more commonly associated with deaths by day 28 after birth. Figures 5 and 6 provide details on perinatal outcomes related to easily recognizable congenital malformations.

**Figure 4:**
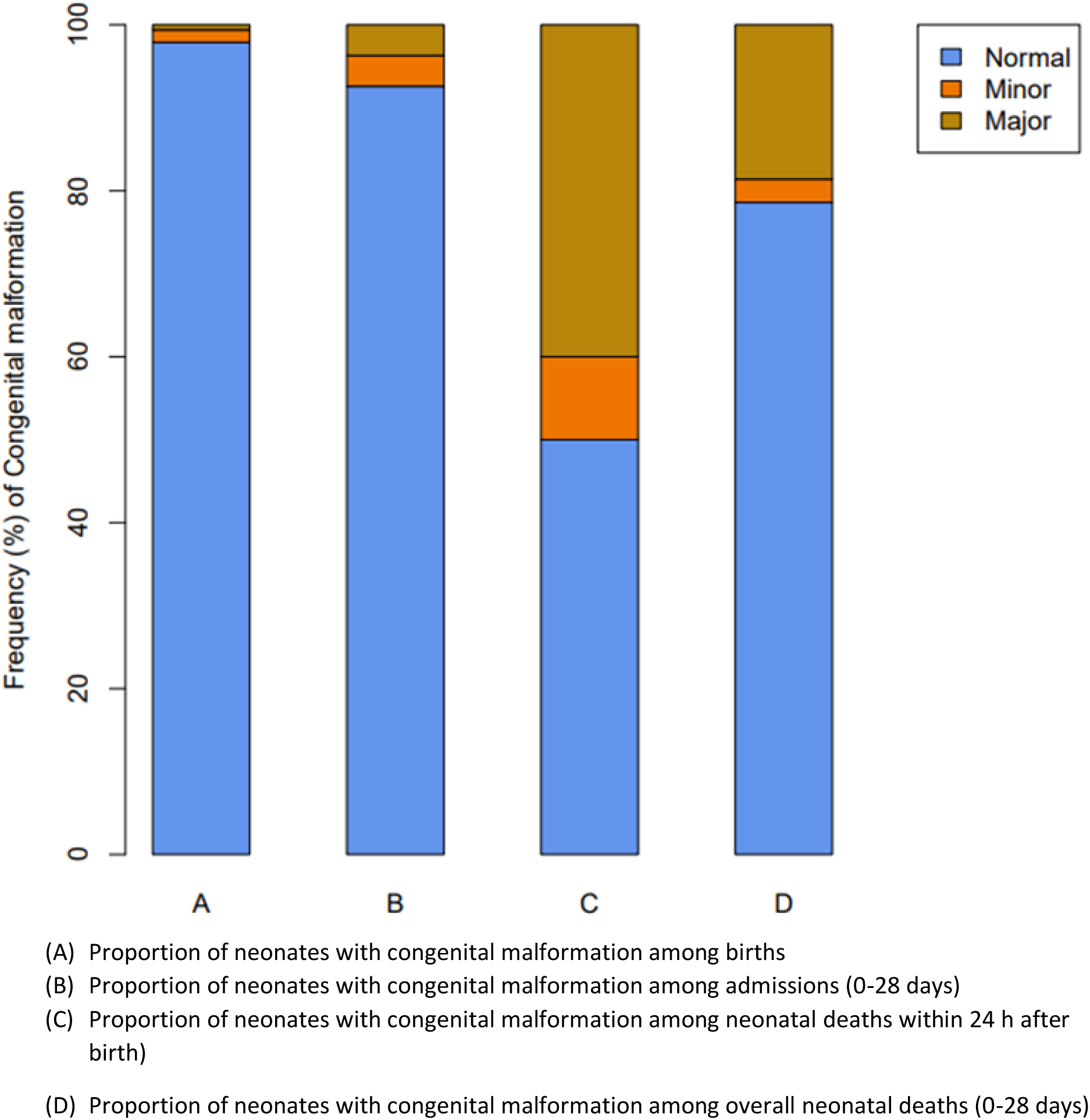
Proportion of neonates with easily recognisable congenital malformations among births, admissions, deaths within 24h and deaths by day 28 in a cohort of Gambian neonates born following relatively healthy pregnancy

**Figure 5:**
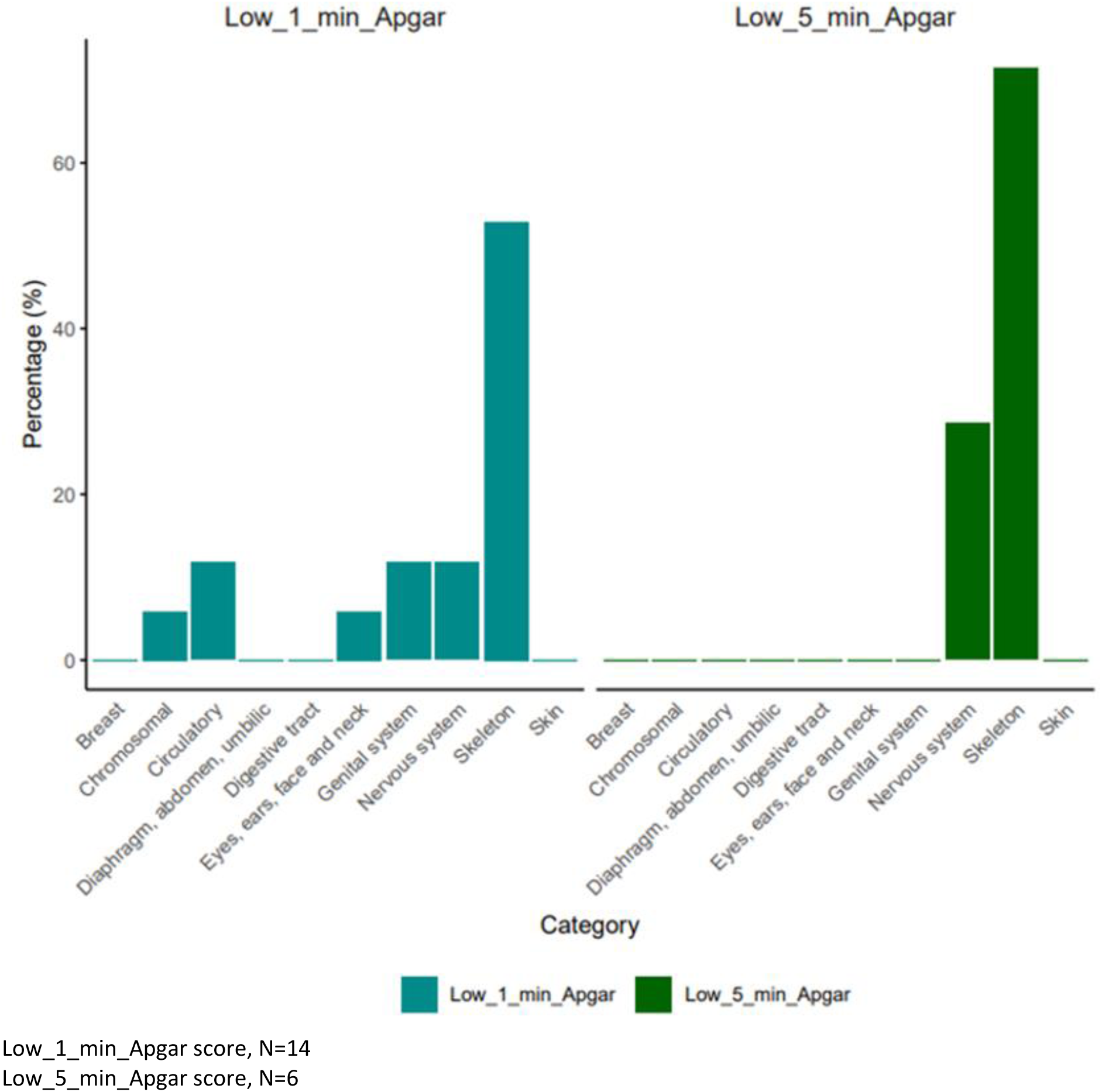
Proportion of easily recognisable congenital malformations among neonates with low Apgar scores (<7)

**Figure 6:**
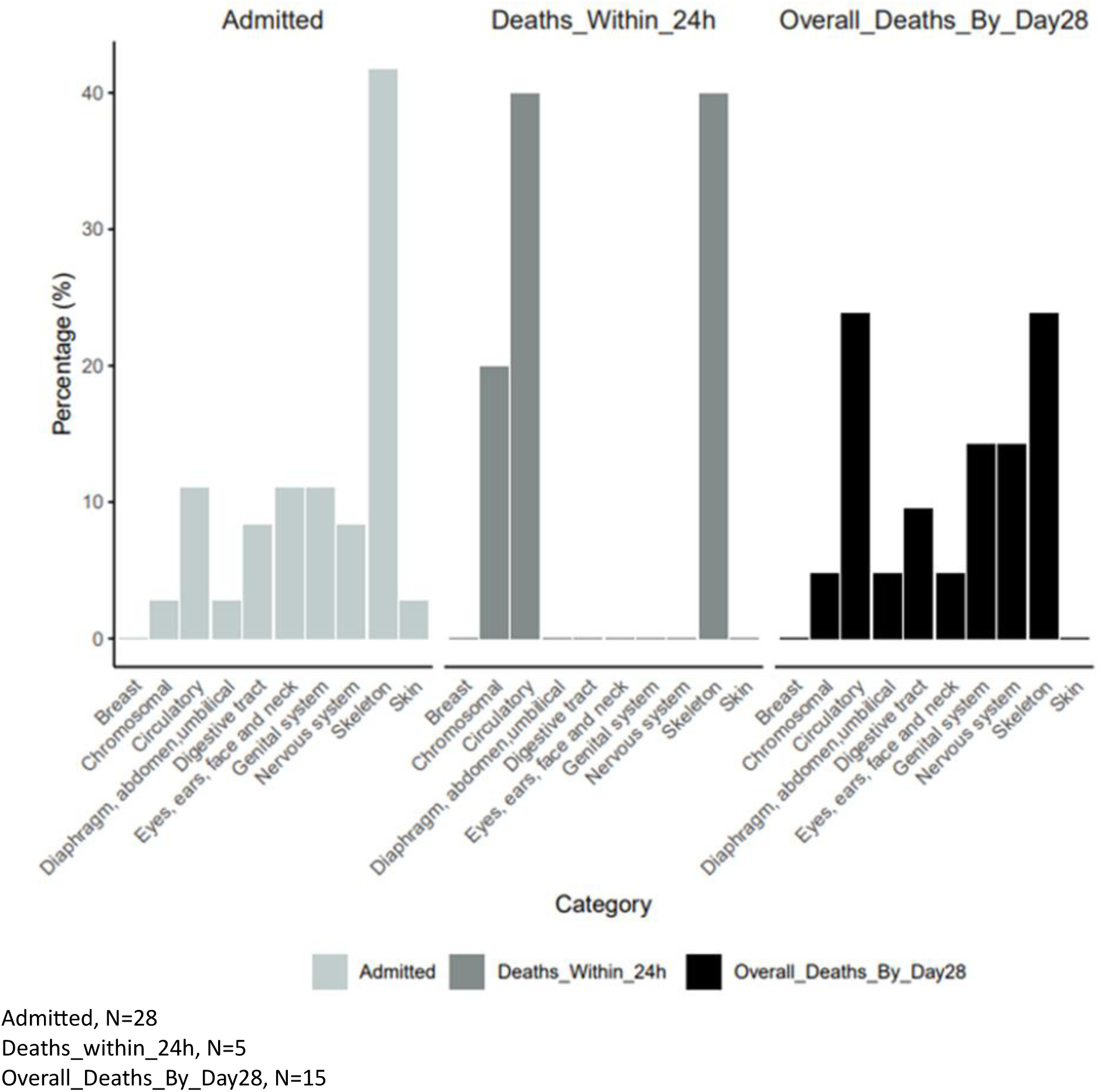
Proportion of each type of easily recognisable congenital malformations among, admissions, deaths within 24h, and deaths by day 28 after birth in a cohort of Gambian neonates

**Table 3:**
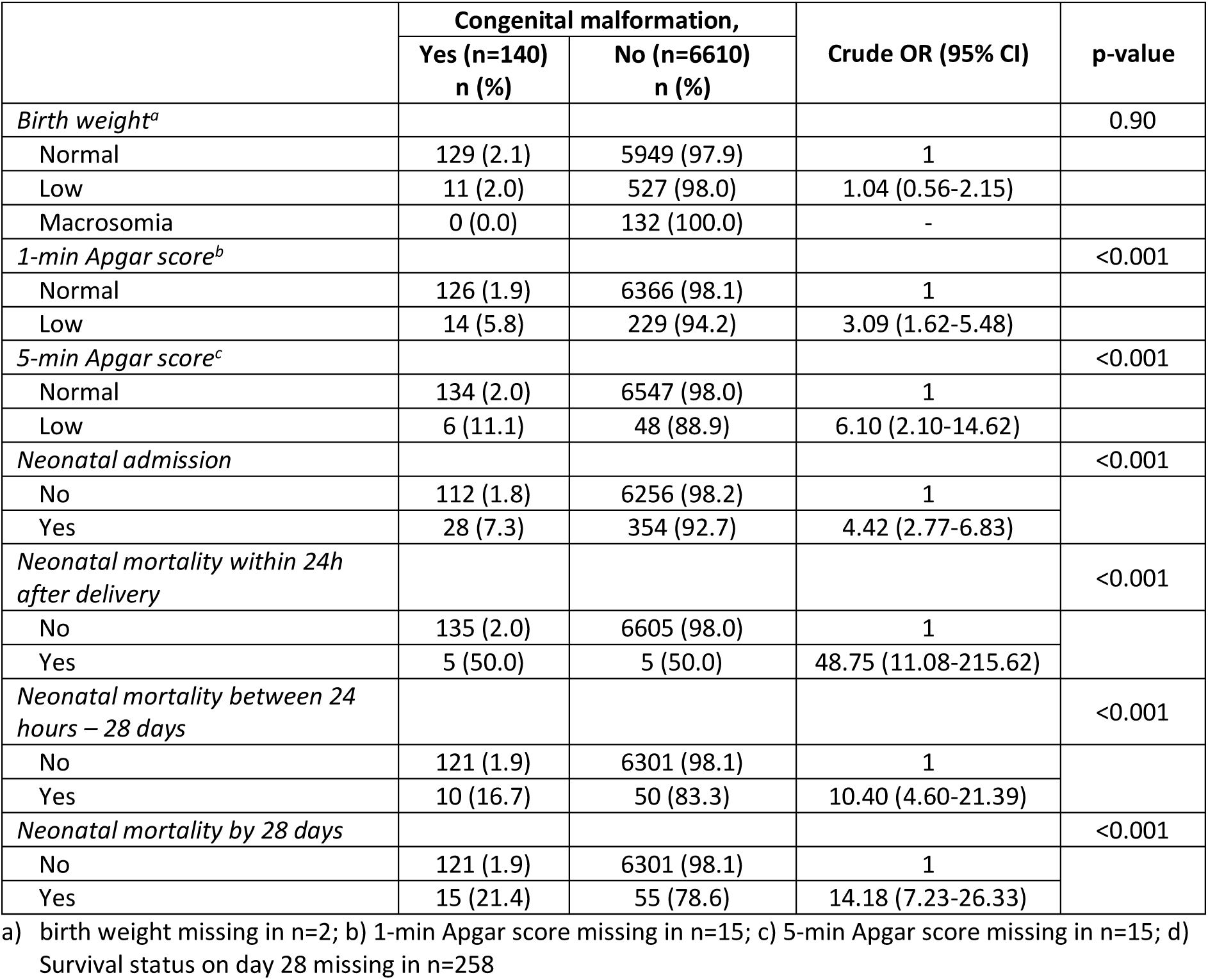
Adverse perinatal outcomes associated with easily recognisable congenital malformations in a cohort of Gambian neonates born following relatively healthy pregnancy

**Table 4:** Adverse perinatal outcomes associated with easily recognisable congenital malformations stratified by degree of congenital malformation in a cohort of Gambian neonates born following relatively healthy pregnancy

|  | Major<br>(n=39) | Normal<br>(n=6610) | OR (95%CI) | p-value | Minor<br>(n=101) | Normal<br>(n=6610) | OR (95%) | p-value |
| --- | --- | --- | --- | --- | --- | --- | --- | --- |
| *Birth weight<sup>a</sup>* |  |  |  |  |  |  |  |  |
| Normal | 34 (0.6) | 5949 (99.4) | 1.65 (0.50-4.29) | 0.29 | 95 (1.6) | 5949 (98.4) | 1.40 (0.62-3.94) | 0.42 |
| Low | 5 (0.9) | 527 (99.1) |  |  | 6 (1.1) | 527 (98.9) |  |  |
| Macrosomia | 0 (0.0) | 132 (100.0) | - |  | 0 (0.0) | 132 (100.0) | - |  |
| *1-min Apgar<sup>b</sup> score* |  |  |  |  |  |  |  |  |
| Normal | 30 (0.5) | 6367 (99.5) |  |  | 96 (1.5) | 6366 (98.5) |  |  |
| Low | 9 (3.8) | 230 (96.2) | 8.33 (3.43-18.27) | <0.001 | 5 (2.1) | 229 (97.9) | 1.45 (0.45-3.54) | 0.42 |
| *5-min Apgar<sup>c</sup> score* |  |  |  |  |  |  |  |  |
| Normal | 36 (0.5) | 6547 (99.5) |  |  | 98 (1.5) | 6547 (98.5) |  | 0.04 |
| Low | 3 (5.9) | 48 (94.1) | 11.35 (2.16-37.88) | <0.001 | 3 (5.9) | 48 (94.1) | 4.17 (0.82-13.30) |  |
| *Neonatal Admission* |  |  |  |  |  |  |  |  |
| No | 25 (0.4) | 6256 (99.6) |  |  | 87 (1.4) | 6256 (98.6) |  |  |
| Yes | 14 (3.8) | 354 (96.2) | 9.89 (4.70-19.97) | <0.001 | 14 (3.8) | 354 (96.2) | 2.84 (1.48-5.09) | <0.001 |
| *Neonatal mortality within 24h* |  |  |  |  |  |  |  |  |
| No | 35 (0.5) | 6605 (99.5) |  |  | 100 (1.5) | 6605 (98.5) |  |  |
| Yes | 4 (44.4) | 5 (55.6) | 148.91 (28.37-728.18) | <0.001 | 1 (16.7) | 5 (83.3) | 13.19 (0.28-119.05) | 0.09 |
| *Neonatal mortality between 24h and 28 days* |  |  |  |  |  |  |  |  |
| No | 24 (0.4) | 6301 (99.6) |  |  | 97 (1.5) | 6301 (98.5) |  |  |
| Yes | 9 (15.3) | 50 (84.7) | 47.06 (18.30-111.63) | <0.001 | 1 (2.0) | 50 (98.0) | 1.30 (0.03-7.75) | 0.80 |
| *Neonatal mortality death on day 28<sup>d</sup>* |  |  |  |  |  |  |  |  |
| No | 24 (0.4) | 6301 (99.6) |  |  | 97 (1.5) | 6301 (98.5) |  |  |
| Yes | 13 (19.1) | 55 (80.9) | 61.81 (27.40-133.95) | <0.001 | 2 (3.5) | 55 (96.5) | 2.36 (0.28-9.18) | 0.22 |

a) birth weight missing in n=2; b) 1-min Apgar score missing in n=15; c) 5-min Apgar score missing in n=15; d) Survival status on day 28 missing in n=258

## Discussion

This large cohort study of “relatively healthy pregnant women” in The Gambia found that 2.1% of newborns had an easily recognisable congenital malformation. Despite this seemingly low prevalence, newborns with malformations constituted more than one in five of all neonatal deaths. Additionally, the presence of these easily recognisable congenital malformations was strongly associated with other adverse perinatal outcomes such as low Apgar scores and hospitalisation, even mild congenital malformations were strongly associated with neonatal hospitalisation. Musculoskeletal congenital malformations such as talipes equinovarus (most common major) and polydactyly (most common minor) were most frequently observed. History of miscarriage was the only identified risk factor for easily recognisable congenital malformations in our cohort.

The observed prevalence of 2.1% of congenital malformations aligns with findings from other West African studies despite notable variations in prevalence across different countries in Sub-Saharan Africa (26) and differences in the methods to define these malformations (27). Notably, within a single country such as Nigeria, prevalence has been reported to range widely, from 1.3% to 6.3% (28). A subgroup analysis for a metanalysis showed a prevalence of 0.7% for congenital malformations in West Africa (29). In our study, easily recognisable congenital malformations most frequently affected the musculoskeletal system, with polydactyly having the highest frequency, followed by talipes equinovarus. Similar observations were made in other resource-limited settings and hospital based studies in Southwestern Nigeria (30), and Uganda (31), where the musculoskeletal system accounted for 43.8%, and 45.7% of malformations, respectively. In all these studies, including ours, the diagnosis of congenital malformations relied on clinical examination. This approach may lead to an over-estimation of the prevalence of musculoskeletal malformations as they are more easily identifiable in the absence of radiological investigations.

Despite the relative low prevalence of easily identifiable congenital malformations among the study newborns, 1 in 2 neonatal deaths within the first 24 hours of life occurred among newborns with congenital malformations, and this extended to 1 in 5 newborns who died within the neonatal period. Our findings, indicating an increased risk of mortality among newborns with easily recognisable congenital malformations, align with a prior, smaller study conducted in the same Gambian region involving a similar population. The earlier study reported 36-fold increased odds of mortality for neonates with major malformation (18). Such strong associations with mortality have also been found in other African studies conducted in Eritrea and Tanzania (32, 33). However, in those studies, congenital malformations were present in 10.1% and 9.0% of neonatal deaths, respectively, compared to our higher figure of 21.4%. These disparities may stem from differences in the study design. The study in Eritrea was a hospital-based retrospective cross-sectional study and focused on admitted newborns, potentially missing deaths among those newborns who died very soon after delivery or were delivered in other health facilities. The study in Tanzania, being registry-based, included exclusively newborns from the neonatal care unit, possibly overlooking early deaths. In the same line, the association of congenital malformation with other poor outcomes such as low-1 min and -5 min Apgar scores, hospitalisation to neonatal intensive care unit, and higher risk of death within first 24h after delivery suggests that for these newborns intra-uterine hypoxia and neonatal encephalopathy may be involved in the causal pathway to mortality (see figure 1). Congenital malformations, especially those of the hip, central nervous system, musculoskeletal system, are associated with breech presentation, (34) which is strongly linked to an increased risk of hypoxic-anoxic events (35) and prolonged labour with potential progression to intra-partum related asphyxia (36) and subsequent neonatal encephalopathy (37). A study conducted in The Netherlands also showed a significant association between a wide variety of congenital malformations and newborn encephalopathy, with the congenital malformation considered to be the probable cause of the encephalopathy in 36% of cases (38). The strong association between congenital malformations and adverse perinatal outcomes observed in our study highlights the importance of antenatal detection of congenital malformations to enable appropriate and timely health facility delivery to avoid intrapartum-related asphyxia, neonatal encephalopathy, and potentially avoidable death. Routine antenatal surveillance using detailed second trimester ultrasound, could improve outcomes by informing delivery planning and timely specialist referrals for these high-risk newborns (39, 40).

A history of miscarriage was associated with congenital malformations in our study. This factor was the only significant risk factor identified in the unadjusted analysis. Similar findings have been reported in studies conducted in low- and middle-income countries. In India, 16.5% of cases of congenital malformations had a history of previous miscarriage (41). Likewise, a study across 72 hospitals in South America revealed an increased risk of congenital malformation in mothers with previous fetal loss (42).. Even though the causal pathway for the association between congenital malformations and previous miscarriage is not fully understood, it has been postulated that spontaneous miscarriage is often correlated with genetic abnormalities incompatible with life (43). Other explanations include recurrent maternal exposures, ranging from nutritional deficiencies (e.g., folic acid), teratogens or infections (e.g. syphilis) that may result in either in-utero demise or disruption of embryogenesis. Therefore, a history of miscarriage could serve as a red flag for potential higher risk of congenital malformation and provide an opportunity for enhanced antenatal care and detailed ultrasound scans for women experiencing recurrent miscarriages. Earlier in-utero detection of congenital malformations would enable family-centered counselling, delivery at an appropriate health facility and prompt postnatal investigation and management, all of which have potential benefits for newborns and their families.

The limitation of our study includes limited access to radiological investigations, which prevented extensive and precise diagnosis of congenital malformations, likely under-estimating the total prevalence, especially for multiple malformations and congenital heart defects; However, our study specifically targeted easily recognisable congenital malformations typically identifiable through clinical examination, such as limb deformities, cleft lip/palate, and neural tube defects. Given this targeted approach, we believe that the potential for detection bias is minimised. However, we acknowledge that we cannot exclude the possibility that some subtle congenital malformations were missed due to human error. For generalisability, future studies should target broader populations and use standardized clinical assessment protocols, including radiological investigations. The inclusion of only relatively healthy pregnant women and their live-born offspring has probably underestimated the prevalence of malformations and further population-based research including both women with complicated pregnancies and stillbirths is needed. However, considering our contextual definition of “relatively healthy women”, we believe our findings contribute to the regional and global literature on this important topic and highlight the important contribution of congenital malformations towards neonatal hospitalisations and mortality. As this was a post-hoc study using an existing dataset, comprehensive data regarding other known risk factors for congenital malformations were not available, e.g., use of medications and nutritional intake during pregnancy or consanguinity between parents. Despite this data limitation, the study’s key findings are scientifically solid, and represent clinical data that is easily ascertained in practice in our setting. The inclusion of only two public health facilities from The Gambia, both situated in urban areas, limits generalisability of our results to the entire country and other rural regions of West Africa. More research is needed to determine prevalence and risk factors in rural areas as differences may exist in nutritional, environmental and genetic exposures.

### Conclusion

Even among relatively healthy women, easily recognisable congenital malformations are associated with substantial neonatal morbidity and mortality and are an important public health issue. Improved antenatal diagnosis of congenital malformations is urgently required to optimise delivery strategies and improve perinatal outcomes, especially for women with a history of previous miscarriage. A national database/register for congenital malformations in the Gambia is also essential for ongoing surveillance/ monitoring and to inform and guide policy. Raising awareness about the importance of regular antenatal visits and early detection of congenital malformations among pregnant women can encourage better use of diagnostic services. Additionally, focused training for healthcare providers on recognising congenital malformations, expanding access to ultrasound screening, and strengthening referral systems to advanced diagnostic centres are key steps toward improving the detection and care of congenital malformations in The Gambia.

## Data Availability

Data may be obtained from a third party and are not publicly available. The clinical data has been collected following provision of informed consent under the prerequisite of strict participant confidentiality. Qualified researchers may request access with the Gambia Government/MRC Joint Ethics Committee. The review process and release of data will be facilitated by MRC Unit The Gambia (http://www.mrc.gm/) through the Head of Governance at MRCG. Access will not be unduly restricted.

## List of abbreviations

BMCHH: Bundung Maternal and Child Health Hospital
DALY: Disability Adjusted Life Years;
eCRF: electronic Case Report Form
EFSTH: Edward Francis Small Teaching Hospital
KGH: Kanifing General Hospital
SDG: Sustainable Development Goal
SHC: Serekunda Health Centre
SSA: Sub-Saharan Africa

## DECLARATIONS

### Ethics approval and consent to participate

This study involved secondary analysis of The PregnAnZI-2 trial. The trial was approved by The Gambia Government/Medical Research Council Unit The Gambia (MRCG) Joint Ethics Committee, the Comité d’Ethique pour la Recherche en Santé (CERS) and the Ministry of Health of Burkina Faso, and the LSHTM Ethics Committee. This included approval for further research utilising trial data. All participants provided written informed consent to participate in the trial including use of their data for further research.

### Competing interests

Dr Usman N. Nakakana reported being an employee of the Gates Foundation in Seattle, United States, and owning shares of the company. No other disclosures were reported.

### Funding

The PregnAnZI-2 trial was funded by a grant from the UKRI under the Joint Global Health Trial Scheme (JGHT) (ref: MC_EX_MR/P006949/1) and support from the Gates Foundation (Ref:OPP1196513). The funders and study sponsor (MRCG at LSHTM) had no role in the study design, collection, analysis, or interpretation of data, writing of article nor the decision to submit for publication.

### Authors’ contributions

AR and UDA conceived and designed the initial trial. HT contributed to the trial design and implementation. SG and NB designed this study with input from AR, HB, and CB. NB performed the analysis with full access to the data and input from CB. SG drafted the initial manuscript with input from NB, HB, AR, CB, and UDA. NB, SG, AR, UDA, HT, HB, CB, BC, UNN, MD, FS, JCJ, ES, SB, OJ, SB, AS, EN, and YN contributed to the writing of the final version, and have read and approved it. AR gave oversight to the work as guarantor and accepts full responsibility for finished work and controlled the decision to publish.

### Consent to publish

Not applicable.

## Acknowledgements

We thank the project managers, Asheme Mahmoud and Bakary Fatty and all the field, data-management, and laboratory teams for providing support to conduct of the main trial. We also thank the BMCHH, SHC, KGH and EFSTH leadership board and staff for their support. We are grateful to all the mothers and their neonates who participated to this study.

## Notes

### Clinical Trial

NCT03199547

### Funding Statement

The PregnAnZI2 trial was funded by a grant from the UKRI under the Joint Global Health Trial Scheme (JGHT) (ref: MC_EX_MR/P006949/1) and support from the Gates Foundation (Ref:OPP1196513). The funders and study sponsor (MRCG at LSHTM) had no role in the study design, collection, analysis, or interpretation of data, writing of article nor the decision to submit for publication.

### Author Declarations

This study involved secondary analysis of The PregnAnZI2 trial. The trial was approved by The Gambia Government Medical Research Council Unit The Gambia (MRCG) Joint Ethics Committee, the Comite de Ethique pour la Recherche en Sante (CERS) and the Ministry of Health of Burkina Faso, and the LSHTM Ethics Committee. This included approval for further research utilising trial data. All participants provided written informed consent to participate in the trial including use of their data for further research.

## References

1. Arnold Christian CPH, Bernadette Modell. Global Report on Birth Defects, The Hidden Toll of dying and disabled children.. March of Dimes Birth Defect Foundation; 2006.

2. CDC. World Birth Defects Day 2022: Global Efforts to Prevent Birth Defects and Support Families Centers for Disease Control and Prevention 2022 [Available from: https://www.cdc.gov/globalhealth/stories/2022/world-birth-defects-day-2022.html.

3. Organization WH, Control CfD, Prevention. Birth defects surveillance: a manual for programme managers. 2020.

4. Ajao AE, Adeoye IA. Prevalence, risk factors and outcome of congenital anomalies among neonatal admissions in OGBOMOSO, Nigeria. BMC Pediatr. 2019;19(1):88.

5. Sachdeva S, Nanda S, Bhalla K, Sachdeva R. Gross congenital malformation at birth in a government hospital. Indian J Public Health. 2014;58(1):54–6.

6. WHO. Birth Defects Fact sheet World Health Organisation 2022 [Available from: https://www.who.int/news-room/fact-sheets/detail/birth-defects.

7. Boyle B, Addor MC, Arriola L, Barisic I, Bianchi F, Csáky-Szunyogh M, et al. Estimating Global Burden of Disease due to congenital anomaly: an analysis of European data. Arch Dis Child Fetal Neonatal Ed. 2018;103(1):F22–f8.

8. Agot GN, Mweu MM, Wang’ombe JK. Prevalence of major external structural birth defects in Kiambu County, Kenya, 2014-2018. Pan Afr Med J. 2020;37:187.

9. GBD Profile: The Gambia Global Burden of diseases, injuries, and risk factors study 2010 Institute for Health Metrics and Evaluation [Available from: https://www.healthdata.org/sites/default/files/files/country_profiles/GBD/ihme_gbd_country_report_the_gambia.pdf.

10. Christianson A, Howson CP, Modell B. March of Dimes: global report on birth defects, the hidden toll of dying and disabled children. March of Dimes: global report on birth defects, the hidden toll of dying and disabled children. 2005.

11. Nelson K, Holmes LB. Malformations due to presumed spontaneous mutations in newborn infants. New England Journal of Medicine. 1989;320(1):19–23.

12. Brent RL. Environmental causes of human congenital malformations: the pediatrician’s role in dealing with these complex clinical problems caused by a multiplicity of environmental and genetic factors. Pediatrics. 2004;113(Supplement_3):957–68.

13. Organization WH. ICD-11 International Classification of Diseases 11th Revision The Global Standard for Diagnostic Health Information 2019 [Available from: https://icd.who.int/en/.

14. Naderi S. Congenital abnormalities in newborns of consanguineous and nonconsanguineous parents. Obstet Gynecol. 1979;53(2):195–9.

15. Aliyu LD. Fetal Congenital Anomalies in Africa: Diagnostic and Management Challenges. 2021 [cited 03.11.2022]. In: Congenital Anomalies in Newborn Infants - Clinical and Etiopathological Perspectives [Internet]. London: IntechOpen, [cited 03.11.2022]. Available from: https://www.intechopen.com/chapters/71783.

16. Abebe S, Gebru G, Amenu D, Mekonnen Z, Dube L. Risk factors associated with congenital anomalies among newborns in southwestern Ethiopia: A case-control study. PLoS One. 2021;16(1):e0245915.

17. Majumder MS, Hess R, Ross R, Piontkivska H. Seasonality of birth defects in West Africa: could congenital Zika syndrome be to blame? F1000Res. 2018;7:159.

18. Bully Camara CO, Reiko Miyahara,. Stillbirths, Neonatal Morbidity, and Mortality in Health Facility Deliveries in Urban Gambia Frontiers in Paediatrics. 2021;9.

19. Bully Camara JDB, Usman N Nakakana et al. Pre-delivery administration of azithromycin to prevent neonatal sepsis and death: a phase iii double-blind randomized clinical trial (PregnAnZI-2 trial) - Protocol. International Journal of Clinical Trials 2022.

20. Roca A, Camara B, Bognini JD, Nakakana UN, Somé AM, Beloum N, et al. Effect of Intrapartum Azithromycin vs placebo on neonatal sepsis and death: a randomized clinical trial. Jama. 2023;329(9):716–24.

21. World Population Prospects: 2022 Revision [Internet]. 2022. Available from: https://population.un.org/wpp/.

22. IGME. Gambia: Neonatal Mortality Rate United Nations Inter-agency Group for Child Mortality Estimation 2021 [Available from: https://childmortality.org/data/Gambia

23. Anya SE, Hydara A, Jaiteh LE. Antenatal care in The Gambia: missed opportunity for information, education and communication. BMC pregnancy and childbirth. 2008;8:1–7.

24. Lyons EJ, Frodsham AJ, Zhang L, Hill AV, Amos W. Consanguinity and susceptibility to infectious diseases in humans. Biology Letters. 2009;5(4):574–6.

25. Camara B, Bognini JD, Nakakana UN, Some AM, Jagne I, Tahita MC, et al. Pre-delivery administration of azithromycin to prevent neonatal sepsis and death: a phase iii double-blind randomized clinical trial (PregnAnZI-2 trial). International Journal of Clinical Trials. 2022;9(1):34-.

26. Adane F, Afework M, Seyoum G, Gebrie A. Prevalence and associated factors of birth defects among newborns in sub-Saharan African countries: a systematic review and meta-analysis. Pan Afr Med J. 2020;36:19.

27. Ritvanen A, Lancaster P, Hausler M, Nelen V, Gillerot Y. World atlas of birth defects. 2003.

28. Ajao AE, Adeoye IA. Prevalence, risk factors and outcome of congenital anomalies among neonatal admissions in OGBOMOSO, Nigeria. BMC pediatrics. 2019;19(1):1–10.

29. Adane F, Afework M, Seyoum G, Gebrie A. Prevalence and associated factors of birth defects among newborns in sub-Saharan African countries: a systematic review and meta-analysis. Pan African Medical Journal. 2020;36(1).

30. Bakare TI, Sowande OA, Adejuyigbe OO, Chinda JY, Usang UE. Epidemiology of external birth defects in neonates in Southwestern Nigeria. Afr J Paediatr Surg. 2009;6(1):28–30.

31. Ochieng J, Kiryowa H, Munabi I, Ibingira C. Prevalence, nature and characteristics of external congenital anomalies at Mulago hospital. East and Central African Journal of Surgery. 2011;16(1).

32. Andegiorgish AK, Andemariam M, Temesghen S, Ogbai L, Ogbe Z, Zeng L. Neonatal mortality and associated factors in the specialized neonatal care unit Asmara, Eritrea. BMC public health. 2020;20:1–9.

33. Mmbaga BT, Lie RT, Olomi R, Mahande MJ, Kvåle G, Daltveit AK. Cause-specific neonatal mortality in a neonatal care unit in Northern Tanzania: a registry based cohort study. BMC pediatrics. 2012;12(1):1–10.

34. Macharey G, Gissler M, Toijonen A, Heinonen S, Seikku L. Congenital anomalies in breech presentation: A nationwide record linkage study. Congenital Anomalies. 2021;61(4):112–7.

35. Hofmeyr GJ, Hannah M, Lawrie TA. Planned caesarean section for term breech delivery. Cochrane Database of Systematic Reviews. 2015(7).

36. Bayih WA, Birhane BM, Belay DM, Ayalew MY, Yitbarek GY, Workie HM, et al. The state of birth asphyxia in Ethiopia: An umbrella review of systematic review and meta-analysis reports, 2020. Heliyon. 2021;7(10):e08128.

37. Dohbit JS, Foumane P, Tochie JN, Mamoudou F, Temgoua MN, Tankeu R, et al. Maternal and neonatal outcomes of vaginal breech delivery for singleton term pregnancies in a carefully selected Cameroonian population: a cohort study. BMJ open. 2017;7(11):e017198.

38. Felix JF, Badawi N, Kurinczuk JJ, Bower C, Keogh JM, Pemberton PJ. Birth defects in children with newborn encephalopathy. Developmental medicine and child neurology. 2000;42(12):803–8.

39. Alanazi AF, Naser AY, Pakan P, Alanazi AF, Alanazi AAA, Alsairafi ZK, et al. Trends of hospital admissions due to congenital anomalies in England and Wales between 1999 and 2019: an ecological study. International Journal of Environmental Research and Public Health. 2021;18(22):11808.

40. Baardman M, du Marchie Sarvaas G, De Walle H, Fleurke-Rozema H, Snijders R, Ebels T, et al. Impact of introduction of 20-week ultrasound scan on prevalence and fetal and neonatal outcomes in cases of selected severe congenital heart defects in The Netherlands. Ultrasound in Obstetrics & Gynecology. 2014;44(1):58–63.

41. Dewangan M, Ali SM, Firdaus U. Pattern of congenital anomalies and risk factors in newborn in a city of a developing country: an observational study. Int J Med Paediat Oncol. 2016;2:152–55.

42. Paz JE, OTANO L, Gadow EC, Castilla EE. Previous miscarriage and stillbirth as risk factors for other unfavourable outcomes in the next pregnancy. BJOG: An International Journal of Obstetrics & Gynaecology. 1992;99(10):808–12.

43. Evidence Indicates Congenital Anomalies Are Main Cause for Spontaneous Abortion. JAMA. 1964;188(11):33-.

